# Health, behavioural, and social correlates of depressive symptoms among Brazilian adults: a preregistered exposure-wide association study with discovery and replication in two independent nationally representative cross-sectional surveys

**DOI:** 10.64898/2026.08.24.26361203

**Authors:** Bruno da Silva Santos, Ives Cavalcante Passos

## Abstract

Depressive disorders are one of the most common psychiatric conditions worldwide. We systematically screened a prespecified exposure panel for associations with depressive symptoms and evaluated cross-wave replication among Brazilian adults. This preregistered exposure-wide association study used independent, nationally representative cross-sectional samples from the 2013 (n=60,202) and 2019 (n=88,531) Brazilian National Health Surveys. 31 general exposures were assessed with survey-weighted regression; four occupational exposures were analysed separately. The primary outcome was a positive Patient Health Questionnaire-9 screen (PHQ-9 >=10); continuous PHQ-9 score was secondary. Discoveries required a Benjamini-Yekutieli-adjusted p<0.05 in 2013; replication required the same coefficient direction and raw p<0.05 in 2019. 21 general exposures were primary discoveries, and all replicated. Associations spanned health status/health care (n=11), behaviour/participation (n=5), and social/material context (n=5). Poor or very poor vs very good self-rated health showed the largest association (adjusted prevalence ratio 10.97, 95% CI 8.79-13.69 in 2013; 12.33, 10.19-14.92 in 2019). Replicated correlates also included morbidity, smoking, prolonged television viewing, diet, group activities, education, income, sanitation, and nearby public space. All 25 continuous-outcome discoveries replicated. All four occupational associations retained the same direction and raw p<0.05 in 2019. This recurrent profile provides a reproducible map for prioritizing longitudinal research but, because both waves were cross-sectional and exposures were modelled separately, does not establish temporality, causality, or independent effects.

## Introduction

Depressive disorders are one of the most common psychiatric conditions worldwide and impose a substantial public health burden in Latin America (Errazuriz et al., 2023). In GBD 2023, Tropical Latin America had the second-highest age-standardised DALY rate for mental disorders among 21 regions; the burden consisted almost entirely of years lived with disability and was led by anxiety disorders and major depressive disorder (GBD 2023 Mental Disorder Collaborators, 2026). Depressive symptoms have been associated with a breadth of behavioural, social, material, health-related, and occupational exposures, usually examined in separate analyses.

Exposure-wide association studies (ExWAS) systematically evaluate a prespecified exposure panel against a common outcome using consistent models and multiplicity control (Patel et al., 2010; VanderWeele et al., 2020). They provide a hypothesis-generating screen of heterogeneous domains, prioritising candidate associations for confirmation while reducing selective emphasis on individually hypothesised findings. Brazil has repeated, independently sampled National Health Surveys (PNS) with harmonisable measures spanning health status, diagnosed conditions, behaviours, participation, and household context. This creates an opportunity to examine which associations recur across survey editions under common definitions and models. In the 2013 and 2019 PNS, Patient Health Questionnaire-9 (PHQ-9)-based symptom screens showed broadly similar sociodemographic and health-related patterning, whereas comparison with self-reported medical diagnosis revealed materially different associations with age, income, education, race or colour, and self-rated health (de Oliveira et al., 2026). Two recent mental-health ExWAS applications used the UK Biobank and a Finnish twin cohort; one confirmed finding in a random split of the same cohort, while the other compared symptom associations across ages within one longitudinal study (Arias-Magnasco et al., 2025; Wang et al., 2023). Neither design tests whether a harmonised association profile recurs across independent national survey editions in a middle-income setting.

In this preregistered study, using the 2013 and 2019 PNS waves, we aimed to (1) systematically identify exposures associated with a positive screen for depressive symptoms (PHQ-9 ≥10) among Brazilian adults in 2013 and estimate the proportion of these discoveries that met the prespecified replication rule in the independent 2019 wave; and (2) repeat the discovery–replication pipeline using the continuous PHQ-9 score, describe replication across prespecified reverse-causation and interpretive domains, and separately examine occupational correlates.

## Methods

### Study design, preregistration, and causal scope

This preregistered ExWAS study used PNS 2013 as the discovery wave and PNS 2019 as the replication wave. Cross-wave replication means that an association satisfied the registered sign-and-p-value rule in the corresponding questionnaire-eligible domain. The protocol and exposure map were registered before any exposure-PHQ-9 model was fitted (OSF: https://osf.io/8nbkv/). Reporting follows the STROBE recommendations for cross-sectional studies (von Elm et al., 2008). The preregistered analytic architecture and the separation of post-outcome analyses are summarized in Supplementary Figure S2.

### Data source and sampling design

Both PNS editions used stratified multistage cluster sampling, with census tracts or groups of tracts as primary sampling units, households as the second stage, and one selected resident per household for the individual questionnaire. The two editions used independent cross-sectional samples; details and differences between their sampling designs have been reported elsewhere (Souza Júnior et al., 2022; Souza-Júnior et al., 2015).

For each wave, the survey design was constructed before adult-domain restriction using UPA_PNS as the primary sampling unit, V0024 as the stratum, and V00291 as the final selected-resident weight; records required complete design variables and positive finite weights. Isolated primary sampling units used the prespecified ‘adjust’ option. The adult domains contained 60,202 records in 2013 and 88,531 in 2019; exposure-specific counts varied with questionnaire routing and complete-case availability (Figure 1).

**Figure 1.**
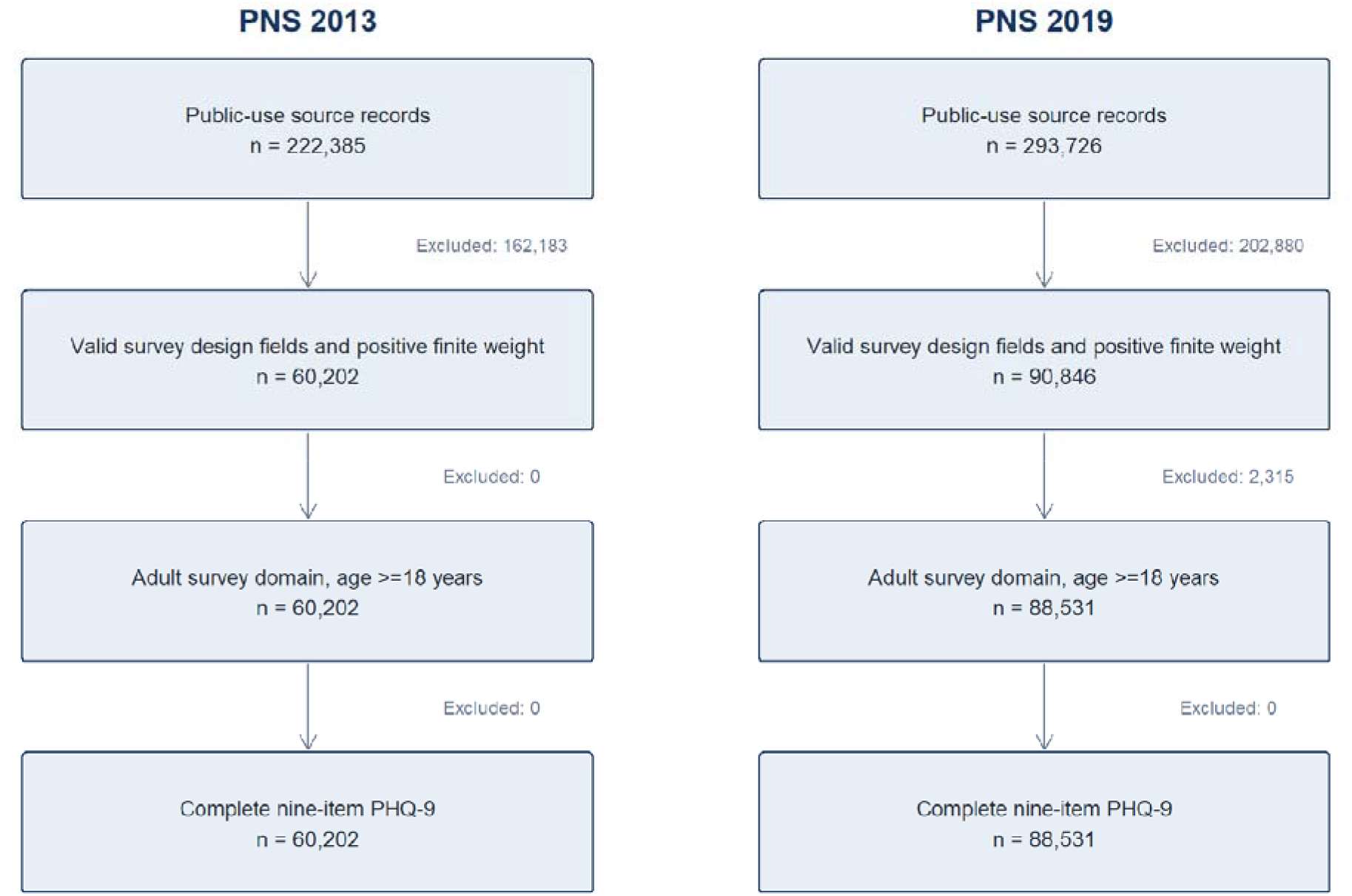
Participant flow in PNS 2013 and PNS 2019. **Legend.** The public-use source files contain records for all household residents; the first transition retains records carrying valid selected-resident design fields and the V00291 individual weight. Selected-resident eligibility began at age 18 in 2013 and age 15 in 2019, after which both waves were restricted to the adult domain (age 18 or older). Counts are unweighted; survey designs were constructed before adult-domain restriction.

### Source population and exposure-specific estimand domains

The source population comprised selected PNS residents aged 18 years or older in each wave. Because questionnaire routing varied, each exposure-PHQ-9 association targeted its prespecified eligible domain rather than one common population. Most general exposures applied to all adults; diabetes and high cholesterol were restricted to ever-tested adults, Family Health Strategy visits to adults in registered households, occupational exposures to currently occupied adults, and passive smoking at work to those in closed or mixed work environments. Complete routing definitions are provided in the registered exposure map.

Accordingly, diabetes and high-cholesterol coefficients describe diagnosis-PHQ-9 associations among ever-tested adults and are not generalized to never-tested adults. Because prior testing may be informative and hypertension was defined among all adults, these cardiometabolic estimates are not directly comparable.

### Outcomes

Depressive symptoms were measured with PHQ-9 items N010-N018. Valid codes 1-4 were shifted to 0-3 by 010: 018 9, yielding 0-27. All nine items had to be valid; there was no prorating or imputation. The primary outcome was PHQ-9 ≥10 and the secondary outcome was the continuous score. In an updated individual-participant-data meta-analysis of 44,503 participants from 100 studies, the ≥10 threshold maximised combined sensitivity and specificity against semi structured diagnostic interviews, with pooled sensitivity and specificity of 0.85 (95% CI 0.79–0.89) and 0.85 (0.82–0.87), respectively (Negeri et al., 2021). Accordingly, PHQ-9 ≥10 was interpreted as a positive depressive-symptom screen rather than a clinical diagnosis.

### Exposure panel and harmonization

The outcome-blind audit retained 35 harmonized candidate correlates: 31 in the general primary family and four in a separate occupational descriptive family. Source variables, valid codes, routing, factor levels, estimand domains, and one target-versus-reference contrast per exposure were prespecified in the registered exposure map; structural non-applicability was not recoded as non-exposure. The retained exposure panel and questionnaire-eligible estimand domains are summarized in Supplementary Table S1.

Before outcome access, each general exposure was assigned to one of three qualitative reverse-causation-risk classes: low (n=9), moderate (n=15), or high (n=7). The rubric considered temporal precedence, external versus symptom-responsive determination, overlap with depressive appraisal or behaviour, and plausible bidirectionality. Each was also assigned to a broad interpretive domain: health status/health care (n=14), behaviour/participation (n=10), or social/material context (n=7). These labels are descriptive aids rather than validated scales or causal taxonomies and cannot be changed after outcomes are opened. The four occupational exposures form the occupational-context domain.

### Covariates

The standard adjustment set comprises age, age squared, sex, self-reported race or colour, educational attainment, within-wave survey-weighted percentile rank of per-capita household income, and geographic region. Age enters linearly and quadratically. Income ranks were derived in the complete adult survey domain before exposure-specific exclusions; ties, including zero income, receive the same weighted rank. To avoid adjusting an exposure for itself, the educational-attainment model omits education and income, and the income model omits income but retains education. No data-driven covariate selection is permitted.

### Statistical analysis

#### Survey models and estimands

All models accounted for strata, clusters, and weights through Taylor-linearized, design-based variance estimation, with each exposure fitted separately in its prespecified estimand domain and complete-case sample. Survey-weighted quasi-Poisson log-link models estimated adjusted prevalence ratios for PHQ-9≥10; Gaussian identity-link models estimated adjusted mean-score differences for continuous PHQ-9. Binary and categorical exposures used prespecified target-versus-reference coefficients. Numeric-score exposures entered as single linear terms, and their displayed contrasts were model-implied rescaled slopes - exp[beta(target - reference)] for the binary outcome and beta(target - reference) for continuous PHQ-9 - which assume linearity on the relevant link scale. Income instead used its prespecified spline contrast. Two-sided 95% confidence intervals and Wald p values used residual survey degrees of freedom.

Binary and categorical exposures used prespecified target-versus-reference contrasts. Ordered quantitative measures used one-degree-of-freedom linear slopes rescaled to the prespecified contrast; nonlinearity diagnostics were reported separately and did not enter the Benjamini-Yekutieli family.

Per-capita household income used a five-knot restricted cubic spline on the within-wave weighted percentile-rank scale, with a 75th-versus-25th-percentile model-matrix contrast. Active-transport minutes used its prespecified 75th-versus-25th-percentile contrast, and the harmful-occupational-agent count used a seven-versus-zero model-implied slope contrast.

### Discovery, multiplicity, and replication

In the binary general family, exactly one frozen primary PNS 2013 p value per exposure was adjusted using the Benjamini-Yekutieli procedure with n=31 and alpha=0.05 (Benjamini and Yekutieli, 2001). The discovery set D comprises exposures with a finite primary PNS 2013 p value and BY-adjusted p<0.05. Only exposures in D were evaluated for general-family replication in PNS 2019. A discovery was considered replicated when its PNS 2019 raw two-sided p value was <0.05 and its primary coefficient had the same sign as in PNS 2013. Direction refers to the log prevalence-ratio coefficient for the binary outcome.

The primary study metric was *R*/|*D*|, where R is the number of discoveries satisfying the replication rule. No binomial confidence interval was calculated because the curated exposure panel is not a random sample of independent trials. Replication p values were not subjected to a second multiplicity correction; raw p<0.05 with the same coefficient sign was the prespecified criterion.

### Secondary and descriptive analyses

The continuous PHQ-9 pipeline independently repeated discovery, BY adjustment with n=31, and replication; its metric was secondary and could not alter the primary binary result. For the binary general family, descriptive summaries reported discovery and replication counts and proportions within the prespecified reverse-causation-risk classes and interpretive domains, without confidence intervals or formal between-stratum comparisons. The four occupational exposures were analysed separately with binary-outcome models in both waves; PNS 2013 p values received BY adjustment with n=4, and wave-specific estimates and replication-status counts were reported without an occupational replication proportion.

### Missing data, diagnostics, and model failures

Analyses used exposure-specific complete cases for the outcome, exposure, and prespecified adjustment set; no multiple imputation was performed. Invalid responses were treated as missing, whereas routing-defined non-applicability determined the estimand domain. Model convergence, warnings, survey degrees of freedom, and coefficient and variance finiteness were checked.

A non-estimable replication did not satisfy the replication rule and was distinguished from an estimable nonsignificant result.

### Post-outcome sensitivity analyses

After completion of the preregistered analysis, supplemental post-outcome protocols evaluated Holm-adjusted replication p values; between-wave coefficient heterogeneity; complete-case availability and outcome distributions; PHQ-9 internal consistency; alternative handling of strata with one contributing primary sampling unit; and departures from the prespecified shapes of ordinal and low-cardinality numeric exposures. These analyses could not reclassify registered discoveries or replications; full methods are provided in the Supplement.

### Software and reproducibility

Analyses used R 4.6.0 and survey 4.5. Pre-outcome integrity checks covered the registered plan, exposure map, microdata, contrasts, BY implementation, and replication rule. Versioned code, package information, and manifests are available in OSF; publicly distributed IBGE microdata are not redistributed.

### Ethics

This secondary analysis used deidentified public-use PNS microdata. The source protocols were approved by the Brazilian National Research Ethics Committee/National Health Council (2013 approval no. 328,159; 2019 approval no. 3,529,376), and source-survey participation was voluntary. No re-identification was attempted.

## Results

### Participants and survey-wave composition

The public-use source files contained 222,385 records in 2013 and 293,726 in 2019. Requiring valid survey-design fields and a positive finite selected-resident weight retained 60,202 and 90,846 records, respectively; adult-domain restriction retained 60,202 in 2013 and 88,531 in 2019. All retained adults had complete nine-item PHQ-9 data (Figure 1).

The weighted mean age was 42.9 years in 2013 and 44.9 years in 2019; women represented 52.9% and 53.2% of the respective samples. Regional distributions were similar, while the weighted proportion identifying as White was 47.6% in 2013 and 43.3% in 2019.

### Outcome distribution and execution integrity

The weighted mean PHQ-9 score was 2.70 in 2013 and 3.42 in 2019; PHQ-9 ≥10 prevalence was 7.9% and 10.8% (Table 1). All adults had nine valid PHQ-9 items. Exposure models then applied exposure- and covariate-specific complete-case criteria; the three records in 2013 and nine in 2019 with missing race or colour were excluded from every adjusted model. The preregistered run completed with 33/33 output hashes verified; all fitted models were estimable and converged without recorded warnings or errors.

**Table 1.** Participant characteristics by PNS wave.

| Characteristic | Category | PNS 2013 | PNS 2019 |
| --- | --- | --- | --- |
| **Participants** | Complete nine-item PHQ-9 | 60,202 | 88,531 |
| **Age, years** | Weighted mean (SD) | 42.9 (16.9) | 44.9 (17.2) |
| **Sex** | Male | 25,920 (47.1%) | 41,662 (46.8%) |
|  | Female | 34,282 (52.9%) | 46,869 (53.2%) |
| **Race or color** | White | 24,106 (47.6%) | 32,409 (43.3%) |
|  | Black | 5,631 (9.1%) | 10,132 (11.5%) |
|  | Asian | 533 (0.9%) | 665 (0.9%) |
|  | Brown/mixed | 29,512 (42.0%) | 44,646 (43.8%) |
|  | Indigenous | 417 (0.4%) | 670 (0.5%) |
| **Educational attainment** | Level 1 (lowest) | 5,807 (8.3%) | 7,632 (6.1%) |
|  | Level 2 | 18,276 (30.7%) | 27,940 (28.7%) |
|  | Level 3 | 5,888 (10.0%) | 6,531 (7.8%) |
|  | Level 4 | 3,327 (5.6%) | 5,474 (6.7%) |
|  | Level 5 | 16,111 (28.1%) | 23,378 (29.8%) |
|  | Level 6 | 3,038 (4.7%) | 3,959 (5.1%) |
|  | Level 7 (highest) | 7,755 (12.7%) | 13,617 (15.8%) |
| **Region** | North | 12,536 (7.5%) | 16,937 (7.8%) |
|  | Northeast | 18,305 (26.5%) | 30,702 (26.5%) |
|  | Southeast | 14,294 (43.9%) | 19,435 (43.4%) |
|  | South | 7,548 (14.8%) | 11,276 (14.7%) |
|  | Central-West | 7,519 (7.4%) | 10,181 (7.6%) |
| **PHQ-9 ≥ 10** | Positive | 5,051 (7.9%) | 9,252 (10.8%) |

**Legend.** Values are unweighted n (survey-weighted %) unless noted; age is the survey-weighted mean (population SD). Race or colour was missing for 3 respondents in 2013 and 9 in 2019 (weighted <0.01% per wave); other listed characteristics were complete. Education levels are the registered standardized cross-wave categories. Percentages may not total 100 because of rounding.

### Primary binary outcome

Twenty-one of the 31 general-family exposures met the PNS 2013 binary discovery rule after BY correction. All 21 had the same coefficient sign and a raw two-sided p value <0.05 in PNS 2019, yielding the preregistered primary metric:

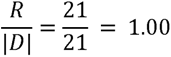

The substantive profile of these replicated associations was not uniform: 11 concerned health status or health care, five behaviour or participation, and five social or material contexts.

Health-status contrasts were the largest. Poor or very poor versus very good self-rated health corresponded to 10.97 times the adjusted PHQ-9-screen prevalence in 2013 (95% CI 8.79-13.69; BY-adjusted p<0.001) and 12.33 times in 2019 (10.19-14.92; raw p<0.001). Poor oral health, heart disease, and chronic spinal problems showed PRs of 2.21-4.19 across waves; hospitalization and the remaining replicated morbidity indicators also had positive associations. Diabetes and high-cholesterol estimates apply only to previously tested adults. Results are reported in full in Table 2.

**Table 2.** Primary binary-outcome discoveries and cross-wave replication.

| ID | Exposure | Frozen contrast | 2013 adjusted PR (95% CI) | 2013 BY p | 2019 adjusted PR (95% CI); raw p |
| --- | --- | --- | --- | --- | --- |
| 4 | Self-rated health | Poor/very poor vs very good | 10.97 (8.79-13.69) | <0.001 | 12.33 (10.19-14.92); p<0.001 |
| 5 | Hospitalization | Yes vs no | 1.92 (1.70-2.16) | <0.001 | 1.95 (1.78-2.12); p<0.001 |
| 7 | Self-rated oral health | Poor/very poor vs very good | 2.96 (2.34-3.76) | <0.001 | 4.19 (3.57-4.91); p<0.001 |
| 8 | Absence of functional dentition (≥13 teeth lost) | Yes vs no | 1.39 (1.22-1.58) | <0.001 | 1.22 (1.11-1.34); p<0.001 |
| 11 | Hypertension | Yes vs no | 1.73 (1.56-1.91) | <0.001 | 1.52 (1.41-1.64); p<0.001 |
| 12 | Diabetes | Yes vs no | 1.53 (1.32-1.77) | <0.001 | 1.61 (1.45-1.78); p<0.001 |
| 13 | High cholesterol | Yes vs no | 1.79 (1.59-2.01) | <0.001 | 1.73 (1.60-1.87); p<0.001 |
| 14 | Heart disease | Yes vs no | 2.84 (2.45-3.28) | <0.001 | 2.21 (2.01-2.43); p<0.001 |
| 16 | Asthma | Yes vs no | 1.91 (1.65-2.21) | <0.001 | 1.91 (1.72-2.12); p<0.001 |
| 17 | Arthritis or rheumatism | Yes vs no | 2.18 (1.93-2.48) | <0.001 | 2.13 (1.94-2.33); p<0.001 |
| 18 | Chronic spinal problem | Yes vs no | 2.55 (2.32-2.79) | <0.001 | 2.29 (2.14-2.45); p<0.001 |
| 27 | Weekly fruit-consumption frequency | 7 vs 0 days/week | 0.82 (0.72-0.94) | 0.024 | 0.72 (0.64-0.80); p<0.001 |
| 28 | Perceived salt consumption | High/very high vs adequate | 1.92 (1.70-2.17) | <0.001 | 1.70 (1.54-1.87); p<0.001 |
| 29 | Group sports or artistic activities | 104 vs 0 occasions/year | 0.66 (0.55-0.80) | <0.001 | 0.70 (0.63-0.77); p<0.001 |
| 33 | Household sanitation classification | Inadequate vs adequate | 0.62 (0.50-0.77) | <0.001 | 0.66 (0.57-0.78); p<0.001 |
| 42 | Educational attainment | Lowest vs highest standardized category | 2.10 (1.70-2.61) | <0.001 | 1.63 (1.40-1.90); p<0.001 |
| 43 | Per-capita household income percentile rank | P75 vs P25 | 0.79 (0.68-0.91) | 0.008 | 0.82 (0.73-0.92); p=0.001 |
| 44 | Current smoking status | Current vs not current | 1.48 (1.33-1.65) | <0.001 | 1.53 (1.40-1.67); p<0.001 |
| 45 | Frequency of smoking inside the household | 365 vs 0 days/year | 1.52 (1.37-1.69) | <0.001 | 1.47 (1.34-1.60); p<0.001 |
| 47 | Television hours per day | ≥6 vs <1 h/day | 1.60 (1.34-1.91) | <0.001 | 1.53 (1.34-1.74); p<0.001 |
| 48 | Public place for physical activity near the residence | Yes vs no | 1.15 (1.04-1.27) | 0.031 | 1.17 (1.08-1.26); p<0.001 |

| ID | Exposure | Frozen contrast | 2013 adjusted PR (95% CI) | 2013 BY p | 2019 adjusted PR (95% CI); raw p |
| --- | --- | --- | --- | --- | --- |

**Table 3.** Descriptive replication summaries by frozen risk class and interpretive domain.

| Stratification | Stratum | Discoveries | Replicated | R/\|D\| |
| --- | --- | --- | --- | --- |
| Reverse-causation risk | High | 3 | 3 | 1.00 |
| Reverse-causation risk | Moderate | 12 | 12 | 1.00 |
| Reverse-causation risk | Low | 6 | 6 | 1.00 |
| Interpretive domain | Health status/health care | 11 | 11 | 1.00 |
| Interpretive domain | Behaviour/participation | 5 | 5 | 1.00 |
| Interpretive domain | Social/material context | 5 | 5 | 1.00 |

**Legend.** No confidence intervals or formal between-stratum comparisons were prespecified. All four occupational exposures were PNS 2013 discoveries after separate BY correction and had the same sign with raw $p < 0.05$ in 2019. Across waves, the model-implied slope-rescaled seven-versus-zero harmful-agent-count contrast yielded PRs of 6.89 and 2.86; PRs were 1.74 and 1.32 for intense physical effort, 1.53 and 1.29 for night work, and 1.62 and 1.82 for passive smoking at work. These are wave-specific descriptive statuses, not an occupational replication proportion.

High or very high perceived salt intake, current smoking, and at least 6 versus less than 1 hour/day of television were associated with 48%-92% higher prevalence across waves (Table 2). Under the registered linear-score models, the slope-rescaled contrasts of seven versus zero fruit-consumption days/week and 104 versus zero group-sport or artistic occasions/year corresponded to 18%-34% lower prevalence (Table 2). These are model-implied contrasts that assume link-scale linearity across the stated ranges.

In the social/material domain, the lowest versus highest standardized education category was associated with PRs of 2.10 in 2013 and 1.63 in 2019; the 75th versus 25th income percentile with PRs of 0.79 and 0.82 (Table 2). The slope-rescaled 365-versus-zero household-smoking-days contrast was positive. Inadequate versus adequate sanitation had PRs of 0.62 and 0.66, while a nearby public place for physical activity had PRs of 1.15 and 1.17 (Table 2).

Ten exposures did not meet the binary discovery rule: employment status, living with a spouse or partner, health-plan coverage, active-transport minutes, association meetings, volunteer work, religious participation, Family Health Strategy visits, endemic-disease-agent visits, and alcohol frequency. Non-discovery under the frozen multiplicity rule is not evidence of no association. Wave-specific estimates for all 31 general-family exposures are shown in Figure 2.

**Figure 2.**
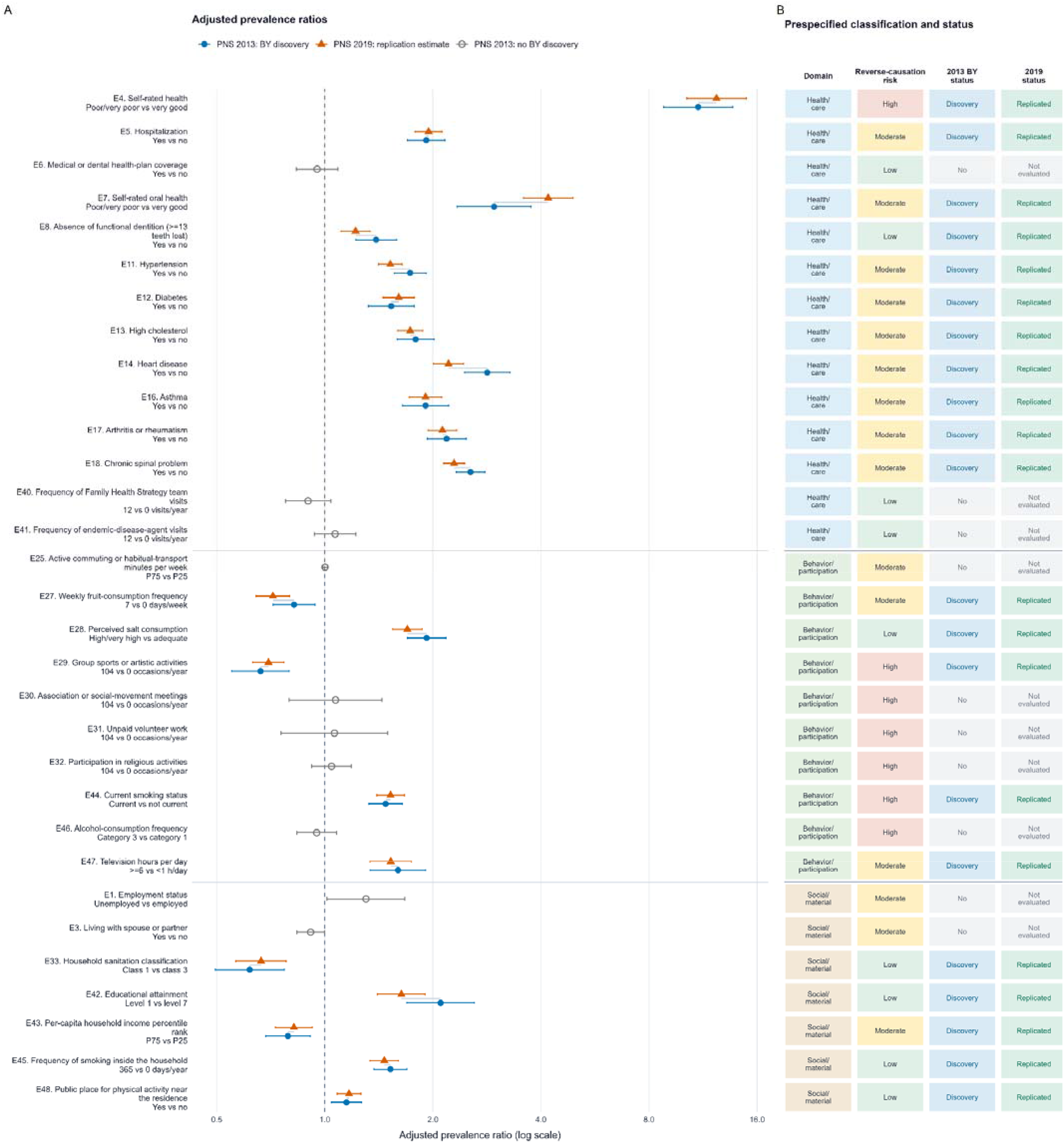
Preregistered primary binary-outcome results across 31 general exposures. **Legend.** Panel A presents adjusted prevalence ratios (PRs) and 95% confidence intervals (CIs) for positive screening for depressive symptoms (PHQ-9 score ≥10) in the independent PNS 2013 and PNS 2019 cross-sectional survey waves. Estimates were obtained using survey-weighted quasi-Poisson regression with a log link and the prespecified exposure-specific adjustment sets and questionnaire-eligible domains. Filled blue circles denote PNS 2013 discoveries after Benjamini–Yekutieli correction across the 31 general exposures; open grey circles denote exposures that did not meet the discovery criterion. Orange triangles show PNS 2019 estimates for the prespecified replication set, and connecting lines link estimates for the same exposure across waves. Panel B shows the frozen interpretive domain, qualitative reverse-causation-risk classification, 2013 discovery status, and 2019 replication status for each exposure. PNS 2019 models were fitted only for the 21 PNS 2013 discoveries; therefore, “Not evaluated” indicates exclusion from the prespecified replication set rather than failure to replicate. Replication required the same coefficient direction as in 2013 and a raw two-sided *p*<0.05 in 2019; all 21 discoveries met this criterion (R/|D|=21/21). Interpretive domains and reverse-causation-risk classifications were specified before outcome-association modelling and are descriptive aids, not validated causal scales or causal taxonomies. Estimates represent associations within exposure-specific eligible populations; replication across independent cross-sectional waves does not establish causal validity or eliminate confounding, reverse causation, selection bias, or measurement differences. The vertical dashed line indicates the null value (PR=1). BY, Benjamini–Yekutieli; CI, confidence interval; PHQ-9, Patient Health Questionnaire-9; PNS, Brazilian National Health Survey; PR, prevalence ratio.

### Secondary continuous outcome

In the separate continuous PHQ-9 pipeline, 25 of 31 exposures met the PNS 2013 discovery rule and all 25 replicated in PNS 2019 (secondary 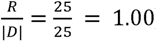). On the 0-27 PHQ-9 scale, adults reporting poor or very poor rather than very good health averaged 6.21 points higher in 2013 (adjusted mean difference 6.21, 95% CI 5.83-6.59; BY-adjusted p<0.001) and 7.30 points higher in 2019 (adjusted mean difference 7.30, 95% CI 6.95-7.65; p<0.001). These are adjusted mean-score differences, not prevalence ratios.

Four exposures discovered only for continuous PHQ-9 also replicated: unemployment (adjusted mean differences 0.69 in 2013 and 0.86 in 2019), living with a spouse or partner (-0.26 and -0.38), association or social-movement meetings (slope-rescaled 104-versus-zero contrast, 0.50 and 0.62), and unpaid volunteer work (corresponding contrast, 0.52 and 0.58); all discovery and replication p values were <0.05. The six continuous non-discoveries were health-plan coverage, active-transport minutes, religious participation, Family Health Strategy visits, endemic-disease-agent visits, and alcohol-consumption frequency. Full results are reported in Supplementary Table S2 and Supplementary Figure S1.

### Stratified and occupational descriptive summaries

The 21 binary discoveries comprised 11 of 14 health/health-care exposures, five of ten behaviour/participation exposures, and five of seven social/material exposures; every discovery in each domain replicated. They also comprised three high-, 12 moderate-, and six low-reverse-causation-risk exposures, with complete replication in each class. These counts were prespecified as descriptive and were not formally compared; replication of a high-risk association does not remove its susceptibility to reverse causation. The occupational descriptive-family estimates are reported in Table 4 and Supplementary Figure S4.

**Table 4.** Occupational descriptive-family results.

| ID | Exposure | Frozen contrast | 2013 adjusted PR<br>(95% CI); BY p | 2019 adjusted PR<br>(95% CI); raw p | Status |
| --- | --- | --- | --- | --- | --- |
| 24 | Intense physical effort at work | Yes vs no | 1.74 (1.50-2.02); BY p<0.001 | 1.32 (1.18-1.48); p<0.001 | Same sign; p<0.05 |
| 36 | Night work | Yes vs no | 1.53 (1.30-1.81); BY p<0.001 | 1.29 (1.13-1.48); p<0.001 | Same sign; p<0.05 |
| 38 | Count of harmful occupational agents (0-7) | 7 vs 0 agents | 6.89 (4.73-10.02); BY p<0.001 | 2.86 (2.15-3.79); p<0.001 | Same sign; p<0.05 |
| 39 | Passive smoking at work | Yes vs no | 1.62 (1.36-1.93); BY p<0.001 | 1.82 (1.57-2.12); p<0.001 | Same sign; p<0.05 |

**Legend.** PR = prevalence ratio; CI = confidence interval; BY = Benjamini-Yekutieli. The ID 38 seven-versus-zero harmful-agent-count PR is a model-implied rescaled slope and assumes link-scale linearity; the other three rows are direct binary contrasts. No occupational replication proportion was prespecified or calculated.

### Post-outcome sensitivity analyses

The sensitivity run completed with 19/19 output hashes verified, and all 46 PNS 2019 models already present in the authoritative primary outputs were reproduced to numerical tolerance. All 21 binary and all 25 continuous replications remained positive under Holm correction of the 2019 replication p values.

None of the 31 binary contrasts showed between-wave heterogeneity after BY correction. Four continuous contrasts did: self-rated health (2019-minus-2013 difference 1.09 PHQ-9 points, 95% CI 0.57 to 1.60; BY p=0.002), self-rated oral health (1.15, 0.67 to 1.64; BY p<0.001), fruit-consumption frequency (-0.45, -0.69 to -0.20; BY p=0.013), and religious participation (-0.36, -0.59 to -0.14; BY p=0.048). The first three were 2013 continuous discoveries and retained their direction with larger absolute contrasts in 2019; religious participation was not a 2013 discovery and changed sign. The four BY-significant continuous contrasts are summarized in Supplementary Table S3, and all between-wave contrasts are shown in Supplementary Figure S3.

No exposure-wave model had a weighted complete-case fraction below 95% (minimum, 97.0%). PHQ-9 distributions differed by hypertension-response completeness, so complete-case selection cannot be excluded. Weighted alpha/omega estimates were 0.845/0.855 in 2013 and 0.859/0.868 in 2019. For the 2013 Family Health Strategy visit model, the prespecified ‘adjust’ and ‘average’ lonely-PSU options produced identical estimates and standard errors. Full results appear in Supplementary Tables S4-S5. Post-outcome nonlinearity diagnostics found Holm-significant departures from rank-linearity for five binary-outcome exposures in 2013, four in 2019, and all seven applicable continuous-outcome exposures in both waves. Saturated models reproduced every frozen extreme contrast; these diagnostics describe category shape and do not revise registered estimates or classifications (Supplementary Table S6).

Among 34 discrete-score shape tests, Holm-significant departures from linearity occurred in 2/8 general binary tests and 5/8 general continuous tests in each wave, and for the occupational harmful-agent count in both waves. For the three score-modelled primary binary discoveries, flexible target-versus-reference PRs preserved direction and were similar to registered estimates (Supplementary Table S7).

The harmful-agent count had sparse upper-tail support and a non-linear pattern in both waves. The registered seven-versus-zero slope-rescaled PR remains the authoritative occupational descriptive estimate but is neither a directly observed endpoint contrast nor evidence of a linear dose-response relationship (Supplementary Table S8).

## Discussion

### Main findings

This preregistered analysis identified a broad and reproducible cross-sectional profile of depressive symptoms in Brazilian adults. Twenty-one of 31 general exposures met the Benjamini-Yekutieli discovery threshold for PHQ-9 ≥10 in PNS 2013, and all 21 satisfied the prespecified replication rule in PNS 2019. The secondary continuous-score analysis produced 25 discoveries, all of which replicated. The binary replicated set was concentrated in health status and health care (11 exposures) but also included behaviour or participation (five) and social or material context (five). Every primary replication remained significant after Holm correction in a post-outcome sensitivity analysis, and no binary coefficient showed multiplicity-adjusted between-wave heterogeneity.

### Health status and health care

The size and consistency of the health-status associations are the main substantive findings. Poor or very poor vs very good self-rated health was associated with an adjusted prevalence of PHQ-9 ≥10 that was 10.97 times higher in 2013 (95% CI 8.79–13.69) and 12.33 times higher in 2019 (95% CI 10.19–14.92), substantially exceeding every other contrast. This pattern is consistent with longitudinal evidence suggesting reciprocal associations between self-rated health and depressive symptoms. Cross-lagged analyses of aging cohorts such as CHARLS and SHARE have found that poorer self-rated health predicts subsequent depressive symptoms, whereas depressive symptoms predict later deterioration, or a lower probability of improvement, in self-rated health (Leng et al., 2025; Liu et al., 2021; Peleg and Nudelman, 2021). In SHARE, the pathway from self-rated health to depression was somewhat stronger overall, particularly among adults aged 65–79 years, whereas the association in the opposite direction was more pronounced among those aged ≥80 years (Peleg and Nudelman, 2021). Similarly, latent growth curve modelling of the Health and Retirement Study found that baseline physical health predicted subsequent depression trajectories and that depressive symptoms predicted physical-health trajectories, further supporting a reciprocal rather than strictly unidirectional relationship (Li et al., 2024). The PNS association is therefore more plausibly interpreted as a strong cross-sectional summary of interrelated perceived health burden and depressive symptomatology than as the effect of a discrete exposure. Because both measures were reported by the same respondent at the same interview, negative affectivity and common-method variance may also have increased the observed magnitude.

Poor self-rated oral health and loss of functional dentition also replicated, although the external evidence is less consistent than for general self-rated health. A meta-analysis of 40 observational studies found a positive association between periodontal disease and depression in case-control studies, with substantial heterogeneity and a need for higher-quality prospective evidence (Zheng et al., 2021). In the UK Biobank, periodontal disease was associated with incident depression, but the prospective estimate was markedly smaller than the cross-sectional estimate (Wang et al., 2024). The PNS estimates, particularly the PR of 4.19 for poor oral health in 2019, may accordingly combine oral disease, pain, functional loss, access to dental care, socioeconomic disadvantage, and symptom-related appraisal.

The replicated morbidity profile was similarly broad: hospitalisation, hypertension, diabetes, high cholesterol, heart disease, asthma, arthritis or rheumatism, and chronic spinal problems were each associated with a higher prevalence of a positive screen. Prior longitudinal and genetically informed work supports reciprocal relationships for some components of this cluster, including cardiovascular disease and type 2 diabetes, although the strength and direction vary across conditions and study designs (Possidente et al., 2023; Wium-Andersen et al., 2020). Hospital use is also plausibly bidirectional. A systematic review found that community depressive symptoms predicted later non-psychiatric admission in unadjusted analyses, but adjusted findings were inconsistent, while a population-based ageing cohort found small increases in subsequent depressive symptoms after hospitalisation or surgery (O’Brien et al., 2018; Prina et al., 2015).

### Behavioural and social patterning

Current smoking, smoking inside the household, at least 6 hours of television per day, and high perceived salt intake were associated with higher positive-screen prevalence, whereas more frequent fruit consumption and group sport or artistic activity were associated with lower prevalence. Lower educational attainment was positively associated with screening, and higher income percentile was inversely associated. Broad lifestyle and socioeconomic patterning has also appeared in large mental-health ExWAS analyses and in PHQ-based analyses of the PNS (Arias-Magnasco et al., 2025; de Oliveira et al., 2026).

The smoking findings are broadly concordant with literature linking tobacco-smoke exposure to depressive symptoms, although the temporally anchored evidence is mixed. In a 20-year Japanese population-based cohort, active smoking alone was not significantly associated with incident depressive symptoms after adjustment (OR 1.27, 95% CI 0.96-1.68), whereas combined active smoking and second-hand smoke exposure was associated with higher odds (ORs 1.39-1.50) (Chu et al., 2026). Cross-sectional NHANES analyses have likewise reported positive associations between tobacco-smoke exposure and depressive symptoms, including a linear relation with serum cotinine and stronger associations among women in one analysis (Fan et al., 2022; Wang et al., 2026). These external data are compatible with, but do not establish, tobacco exposure as an antecedent of depressive symptoms; reverse causation remains plausible because depressive symptoms may influence smoking behaviour or time spent in smoking environments.

For sedentary behaviour, the prospective evidence base is stronger. Meta-analyses of prospective studies consistently distinguish mentally passive sedentary behaviour such as television viewing from mentally active sedentary behaviour such as computer use: television viewing has been associated with higher subsequent depression risk, including dose-response patterns (Huang et al., 2020; Zhou et al., 2023). This literature supports treating the replicated PNS television association as a priority for longitudinal investigation.

For diet, prospective cohort evidence is more consistent for fruit intake, whereas vegetable associations vary across populations and cohort composition (Zamanian et al., 2026). A longitudinal twin analysis also used co-twin comparisons to address shared familial and genetic confounding, although the evidence for vegetables remained less uniform (Matison et al., 2024). In adults aged 45 years or older, a separate meta-analysis associated higher Dietary Inflammatory Index scores and Western dietary patterns with incident depression, while Mediterranean and broadly healthy dietary patterns were not significantly associated (Matison et al., 2021). The PNS fruit-frequency result is therefore concordant with prospective evidence for fruit intake, but it should not be generalized to overall diet quality.

### What replication does and does not establish

The complete replication proportion requires a bounded interpretation. Its denominator comprised associations selected because they had already passed a stringent discovery threshold in 60,202 adults; it is not a random sample of hypotheses and does not estimate the probability that an arbitrary exposure-depression association is true. PNS 2019 also had a larger adult sample and a higher positive-screen prevalence than PNS 2013, which increased power to satisfy a p-value-based rule. Finally, the rule required agreement of sign and statistical threshold, not equivalence of effect size. Four continuous contrasts showed between-wave heterogeneity despite retaining the registered replication classification for the three that were discoveries. Thus, 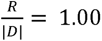 describes threshold-based reproducibility of this selected panel within two PNS editions.

The two directionally counterintuitive findings make this limit concrete. Inadequate sanitation was associated with lower positive-screen prevalence (PR 0.62 in 2013 and 0.66 in 2019), whereas a nearby public place for physical activity was associated with higher prevalence (PR 1.15 and 1.17). Neither direction corresponds to a simple environmental intervention effect. In the Northern Finland Birth Cohort 1966, higher population density and urbanicity were associated cross-sectionally with more severe depressive symptoms, whereas greater residential greenness was associated with less severe symptoms after adjustment for physical activity and individual covariates (Rautio et al., 2024). Conversely, a two-city Chinese cross-sectional study found that better perceived street environments and greater sports-facility density were associated with fewer depressive symptoms, with city-specific patterns for other neighbourhood dimensions (Zhang et al., 2025).

Further evidence shows why recurrence alone cannot resolve contextual confounding or selection. In nationally representative Irish data, area-level deprivation was not independently associated with mental-health outcomes after individual socioeconomic and health adjustment, although perceived safety, cleanliness, service provision, and social-group participation remained associated (Mohan and Barlow, 2023). In Rotterdam, an association between neighbourhood income and reimbursed antidepressant or antipsychotic medication was strongly attenuated after modelling residential self-selection, while the association with neighbour contact emerged only after selection adjustment (Boderie et al., 2023). Among older adults in Ghana and South Africa, the composition and levels of social capital differed by setting, and associations with depression varied by country and were not uniformly beneficial (Adjaye-Gbewonyo et al., 2019). In the PNS models, urbanicity, neighbourhood composition, and residential selection were not included in the preregistered adjustment set. Neither counterintuitive result supports a protective effect of inadequate sanitation or a harmful effect of public recreational space; their recurrence under the same rule as the clinically plausible findings shows that replication can reproduce stable confounding.

### Occupational findings

The occupational family was intentionally separate and should remain descriptive. All four occupational coefficients had the same sign and raw p < 0.05 in 2019, but no occupational replication proportion was prespecified. The contrast for the count of harmful occupational agents attenuated from PR 6.89 to 2.86, relied on very sparse upper-tail observations, and failed the assumed linear-score shape diagnostic in both waves. It is a model-implied seven-versus-zero contrast, not a directly observed endpoint comparison or evidence of a dose-response gradient. Differences between the occupational questionnaires and the composition of the employed population may also have contributed to the attenuation.

### Strengths and limitations

The study’s main strengths are the preregistration, outcome-blind harmonisation, frozen contrasts and estimand domains, design-based estimation, multiplicity control for dependent tests, and evaluation in an independent national survey wave. Versioned post-outcome diagnostics were kept separate from the registered analysis and could not change discovery or replication status. These safeguards reduce selective analytical flexibility and make the reported recurrence auditable.

Several limitations determine what the findings support. Both surveys are cross-sectional, so exposure and symptom timing cannot be established. The panel comprises questionnaire measures that could be harmonised across PNS editions and is not a complete exposome. Most variables were self-reported, and several appraisals share measurement modality with the PHQ-9. Exposures were modelled separately; estimates are marginal associations adjusted for a common covariate set, not mutually independent contributions. Education and income used exposure-specific adjustment sets and should not be compared as if they shared one specification. Complete-case fractions exceeded 95%, but outcome distributions differed by hypertension-response completeness, leaving possible selection. Diabetes and high-cholesterol estimates apply only to previously tested adults. The outcome is a symptom-screening threshold rather than a diagnosis. This distinction is consequential in the PNS: de Oliveira et al. (2026) found close agreement between PHQ-8 and PHQ-9 symptom screens, but substantially different sociodemographic patterning when symptom-based measures were compared with self-reported medical diagnosis. Relative to PHQ-9 screening, associations with formal diagnosis were weaker among Black and Brown adults and those reporting fair or poor health, but stronger at older ages and higher socioeconomic position. Thus, the associations reported here describe the distribution of depressive symptom burden rather than the distribution of clinically recognised depressive disorder.

The replicated profile may help prioritise longitudinal questions rather than nominate immediate preventive targets. Future analyses should separate temporal pathways within the dominant health-status cluster, using repeated symptom measures, objective disease ascertainment where available, and explicit modelling of pain, disability, health-care contact, and urban context.

## Conclusion

Within a nationally representative Brazilian framework, this preregistered screen identified 21 associations with a positive PHQ-9 screen that met a stringent multiplicity-corrected threshold in 2013, and all 21 recurred in an independent 2019 sample under a replication rule fixed in advance. The replicated set is dominated by health-status appraisals but extends to behaviour, participation and social and material conditions. The contribution is a transparent, auditable map of which prespecified cross-sectional associations recur across the two survey editions.

## Declarations

### Author contributions

**BdSS**: Conceptualization, Data Curation, Formal Analysis, Investigation, Visualization, Writing – original draft, Writing – review & editing. **ICP**: Supervision, Validation, Writing – review & editing.

## Data and code availability

The preregistration, frozen plan, exposure map, and integrity manifests are publicly available at https://osf.io/8nbkv/. Versioned post-outcome sensitivity and shape-diagnostic protocols, runners, and non-disclosive derived outputs are available in the same OSF project. PNS public-use microdata are distributed by IBGE and are not redistributed by the authors.

## Declaration of generative AI and AI-assisted technologies in the manuscript preparation process

During the preparation of this work, the authors used OpenAI’s ChatGPT (5.6 Sol, https://chatgpt.com/) and Anthropic’s Claude (Opus 5, https://claude.ai) as AI-assisted writing tools to support readability of the main text and to aid in the treatment of code. The authors reviewed and edited the content as needed and take full responsibility for the published article.

