## Supplementary for "Health, behavioural, and social correlates of depressive symptoms among Brazilian adults: a preregistered exposure-wide association study with discovery and replication in two independent nationally representative cross-sectional surveys"

0000-0001-6407-8219

*^a^ Laboratory of Molecular Psychiatry, Experimental Research Center (CPE) and Clinical Research Center (CPC), Hospital de Clínicas de Porto Alegre (HCPA), Porto Alegre, RS, Brazil.*

*^b^ Federal University of Rio Grande do Sul (UFRGS), School of Medicine, Graduate Program in Psychiatry and Behavioral Sciences, Department of Psychiatry, Porto Alegre, RS, Brazil.*

*^c^ National Institute of Science and Technology in Digital Mental Health (INCT-SMD), Rio Grande do Sul, Porto Alegre, Brazil.*

**Supplementary Methods	3**

Post-outcome sensitivity analyses	3

Table S1. Frozen exposure panel and questionnaire-eligible estimand domains	3

Supplementary Results	6

Table S2. Continuous PHQ-9 discoveries and cross-wave replication	6

Post-outcome sensitivity results	7

Replication and between-wave heterogeneity	7

Table S3. Continuous contrasts with BY-adjusted between-wave heterogeneity	7

Missingness, internal consistency, and survey-design diagnostics	8

Table S4. Exposure-wave rows with weighted complete-case availability below 99%	8

Table S5. Descriptive PHQ-9 internal consistency by wave	8

Exposure-shape diagnostics	8

Table S6. Post-outcome ordinal nonlinearity diagnostic summary	8

Table S7. Post-outcome discrete-score shape diagnostic summary	9

Table S8. Support for the occupational harmful-agent count (ID 38)	10

**Supplementary Figures	11**

Figure S1. Secondary continuous PHQ-9 results across 31 general exposures.	11

Figure S2. Preregistered discovery-replication architecture. Solid boxes denote the frozen pre-outcome analysis; the dashed box contains post-outcome sensitivity analyses.	12

Figure S3. Post-outcome between-wave heterogeneity sensitivity across all 31 general exposures.	13

Figure S4. Occupational descriptive-family results in employed or workplace-eligible adults.	14

### Supplementary Methods

Figure S2 summarizes the separation between the frozen preregistered workflow and the post-outcome sensitivity analyses. Table S1 lists the retained exposure panel and its questionnaire-eligible estimand domains.

#### Post-outcome sensitivity analyses

After the preregistered run was completed, an explicitly post-outcome supplemental protocol was frozen before these additional analyses were executed. These analyses could not reclassify the registered discoveries or replications. Within each frozen discovery set, PNS 2019 replication p values were adjusted by Holm. Coefficient heterogeneity was evaluated for all 31 general exposures as beta_2019 - beta_2013, using the sum of the independent wave-specific variances and BY correction across 31 tests. The active-transport contrast was rescaled to a common 60-minutes-per-week unit. A nonsignificant heterogeneity test was not interpreted as evidence of equivalence.

The supplemental diagnostics also quantified exposure- and covariate-specific complete-case availability, weighted PHQ-9 distributions among complete and incomplete records, and analytic strata with one contributing PSU. PHQ-9 internal consistency was summarized separately by wave using alpha from the survey-weighted covariance matrix, a one-factor omega approximation from the weighted Pearson correlation matrix, and weighted item-rest correlations. These are descriptive internal-consistency measures, not test-retest reliability. Models triggering the lonely-PSU diagnostic were repeated with survey.lonely.psu = 'average' and compared with the registered 'adjust' specification.

A second, separately versioned post-outcome diagnostic examined the eight general-family exposures frozen as ordinal categorical variables (IDs 3, 4, 7, 28, 33, 42, 46, and 47). For K ordered categories, the saturated factor model was re-expressed with orthogonal polynomials on equally spaced category ranks and the K-2 higher-order terms were tested jointly. Holm adjustment was applied separately within each wave-by-outcome family of seven applicable tests; ID 3 had only two categories. Category rank was only a diagnostic coding device, not an assumption of substantive equal spacing or dose response. All fitted models had to reproduce the authoritative frozen target-versus-reference contrast.

A third, separately versioned post-outcome diagnostic examined eight low-cardinality general-family numeric-score exposures (IDs 27, 29-32, 40, 41, and 45) and the occupational harmful-agent count (ID 38). For each observed score support, a saturated polynomial basis evaluated at the actual numeric values replaced the registered single linear term; the higher-order terms were tested jointly for departure from the registered link-scale shape. Holm correction was applied separately within each wave-by-outcome-by-family set. Every registered slope refit had to reproduce the authoritative estimate, and flexible target-versus-reference contrasts and level-specific support were descriptive only. ID 25 was excluded from this diagnostic because of its high-cardinality minute scale, while ID 43 already used a prespecified spline.

| Table S1. Frozen exposure panel and questionnaire-eligible estimand domains | | | | |
| --- | --- | --- | --- | --- |
| **ID** | **Exposure** | **Family** | **Domain** | **Estimand population** |
| 1 | Employment status | General | Social/material | All adults |
| 3 | Living with spouse or partner | General | Social/material | All adults |
| 4 | Self-rated health | General | Health/health care | All adults |
| 5 | Hospitalization | General | Health/health care | All adults |
| 6 | Medical or dental health-plan coverage | General | Health/health care | All adults |
| 7 | Self-rated oral health | General | Health/health care | All adults |
| 8 | Absence of functional dentition (>=13 teeth lost) | General | Health/health care | All adults |
| 11 | Hypertension | General | Health/health care | All adults |
| 12 | Diabetes | General | Health/health care | Ever glucose-tested adults |
| 13 | High cholesterol | General | Health/health care | Ever cholesterol-tested adults |
| 14 | Heart disease | General | Health/health care | All adults |
| 16 | Asthma | General | Health/health care | All adults |
| 17 | Arthritis or rheumatism | General | Health/health care | All adults |
| 18 | Chronic spinal problem | General | Health/health care | All adults |
| 24 | Intense physical effort at work | Occupational | Occupational | Currently occupied adults |
| 25 | Active commuting or habitual-transport minutes per week | General | Behavior/participation | All adults |
| 27 | Weekly fruit-consumption frequency | General | Behavior/participation | All adults |
| 28 | Perceived salt consumption | General | Behavior/participation | All adults |
| 29 | Group sports or artistic activities | General | Behavior/participation | All adults |
| 30 | Association or social-movement meetings | General | Behavior/participation | All adults |
| 31 | Unpaid volunteer work | General | Behavior/participation | All adults |
| 32 | Participation in religious activities | General | Behavior/participation | All adults |
| 33 | Household sanitation classification | General | Social/material | All adults |
| 36 | Night work | Occupational | Occupational | Currently occupied adults |
| 38 | Count of harmful occupational agents (0-7) | Occupational | Occupational | Currently occupied adults |
| 39 | Passive smoking at work | Occupational | Occupational | Occupied adults in closed/mixed work environments |
| 40 | Frequency of Family Health Strategy team visits | General | Health/health care | Adults in ESF-registered households |
| 41 | Frequency of endemic-disease-agent visits | General | Health/health care | All adults, regardless of ESF registration |
| 42 | Educational attainment | General | Social/material | All adults |
| 43 | Per-capita household income percentile rank | General | Social/material | All adults |
| 44 | Current smoking status | General | Behavior/participation | All adults |
| 45 | Frequency of smoking inside the household | General | Social/material | All adults |
| 46 | Alcohol-consumption frequency | General | Behavior/participation | All adults |
| 47 | Television hours per day | General | Behavior/participation | All adults |
| 48 | Public place for physical activity near the residence | General | Social/material | All adults |
| **Legend**. ESF = Family Health Strategy. Labels are concise English descriptors; the registered JSON map is authoritative for wave-specific source variables, coding expressions, target/reference contrasts, and structural-missingness rules. General-family IDs 25, 27, 29-32, 40, 41, and 45 and occupational ID 38 are modeled as single linear numeric terms; their displayed target-versus-reference quantities are model-implied rescaled slopes rather than direct category contrasts. | | | | |

**Reverse-causation-risk classes (general family only).** Low: IDs 6, 8, 28, 33, 40, 41, 42, 45, and 48; moderate: IDs 1, 3, 5, 7, 11-14, 16-18, 25, 27, 43, and 47; high: IDs 4, 29-32, 44, and 46. These qualitative classes were frozen before outcome access and were used only for descriptive stratification.

**Outcome-blind exclusions.** Candidate measures were not retained when they failed cross-wave harmonization, overlapped temporally or conceptually with PHQ-9, duplicated a retained construct, lacked a 2019 counterpart, fell outside the prespecified panels, or had unresolved comparability. The item-level exclusion inventory is archived in the preregistration bundle; retained IDs therefore remain intentionally nonconsecutive.

### Supplementary Results

| Table S2. Continuous PHQ-9 discoveries and cross-wave replication | | | | | |
| --- | --- | --- | --- | --- | --- |
| **ID** | **Exposure** | **Frozen contrast** | **2013 adjusted mean difference (95% CI)** | **2013 BY p** | **2019 adjusted mean difference (95% CI); raw p** |
| 1 | Employment status | Unemployed vs employed | 0.69 (0.34-1.03) | <0.001 | 0.86 (0.55-1.16); p<0.001 |
| 3 | Living with spouse or partner | Yes vs no | -0.26 (-0.38--0.13) | <0.001 | -0.38 (-0.50--0.26); p<0.001 |
| 4 | Self-rated health | Poor/very poor vs very good | 6.21 (5.83-6.59) | <0.001 | 7.30 (6.95-7.65); p<0.001 |
| 5 | Hospitalization | Yes vs no | 1.60 (1.33-1.86) | <0.001 | 2.12 (1.85-2.39); p<0.001 |
| 7 | Self-rated oral health | Poor/very poor vs very good | 2.61 (2.25-2.97) | <0.001 | 3.76 (3.44-4.09); p<0.001 |
| 8 | Absence of functional dentition (>=13 teeth lost) | Yes vs no | 0.84 (0.64-1.04) | <0.001 | 0.55 (0.37-0.72); p<0.001 |
| 11 | Hypertension | Yes vs no | 1.24 (1.07-1.41) | <0.001 | 1.07 (0.92-1.22); p<0.001 |
| 12 | Diabetes | Yes vs no | 1.13 (0.81-1.45) | <0.001 | 1.29 (1.06-1.52); p<0.001 |
| 13 | High cholesterol | Yes vs no | 1.50 (1.28-1.72) | <0.001 | 1.70 (1.51-1.89); p<0.001 |
| 14 | Heart disease | Yes vs no | 2.96 (2.52-3.40) | <0.001 | 2.73 (2.41-3.04); p<0.001 |
| 16 | Asthma | Yes vs no | 1.79 (1.47-2.11) | <0.001 | 1.99 (1.68-2.30); p<0.001 |
| 17 | Arthritis or rheumatism | Yes vs no | 2.42 (2.10-2.74) | <0.001 | 2.58 (2.30-2.86); p<0.001 |
| 18 | Chronic spinal problem | Yes vs no | 2.38 (2.21-2.55) | <0.001 | 2.38 (2.22-2.54); p<0.001 |
| 27 | Weekly fruit-consumption frequency | 7 vs 0 days/week | -0.35 (-0.51--0.18) | <0.001 | -0.79 (-0.97--0.61); p<0.001 |
| 28 | Perceived salt consumption | High/very high vs adequate | 1.23 (1.05-1.42) | <0.001 | 1.34 (1.15-1.53); p<0.001 |
| 29 | Group sports or artistic activities | 104 vs 0 occasions/year | -0.37 (-0.53--0.20) | <0.001 | -0.64 (-0.78--0.50); p<0.001 |
| 30 | Association or social-movement meetings | 104 vs 0 occasions/year | 0.50 (0.18-0.82) | 0.011 | 0.62 (0.21-1.03); p=0.003 |
| 31 | Unpaid volunteer work | 104 vs 0 occasions/year | 0.52 (0.14-0.90) | 0.034 | 0.58 (0.22-0.94); p=0.002 |
| 33 | Household sanitation classification | Inadequate vs adequate | -0.75 (-0.99--0.51) | <0.001 | -0.86 (-1.07--0.65); p<0.001 |
| 42 | Educational attainment | Lowest vs highest standardized category | 1.14 (0.84-1.43) | <0.001 | 0.95 (0.68-1.22); p<0.001 |
| 43 | Per-capita household income percentile rank | P75 vs P25 | -0.44 (-0.61--0.26) | <0.001 | -0.45 (-0.63--0.26); p<0.001 |
| 44 | Current smoking status | Current vs not current | 0.79 (0.61-0.97) | <0.001 | 0.98 (0.80-1.15); p<0.001 |
| 45 | Frequency of smoking inside the household | 365 vs 0 days/year | 0.98 (0.79-1.18) | <0.001 | 1.07 (0.88-1.26); p<0.001 |
| 47 | Television hours per day | >=6 vs <1 h/day | 1.12 (0.76-1.49) | <0.001 | 1.33 (1.03-1.63); p<0.001 |
| 48 | Public place for physical activity near the residence | Yes vs no | 0.31 (0.18-0.45) | <0.001 | 0.48 (0.36-0.61); p<0.001 |
| **Legend**. CI = confidence interval; BY = Benjamini-Yekutieli. The table contains only PNS 2013 continuous-outcome discoveries; every row satisfied the frozen PNS 2019 replication rule. For IDs 27, 29, 30, 31, and 45, displayed target-versus-reference differences are model-implied rescaled slopes and assume identity-link linearity across the stated range; other rows use direct category contrasts except income (ID 43), which uses the frozen spline contrast. Sanitation is labeled by meaning. The education model omits education and income; the income model omits income but retains education. Complete paired continuous-outcome estimates are visualized in Figure S1. | | | | | |

#### Post-outcome sensitivity results

##### Replication and between-wave heterogeneity

**Holm-adjusted replication.** All 21 binary and all 25 continuous registered replications remained positive after adjustment of PNS 2019 replication p values within their respective frozen discovery sets. Exposure-level adjusted p values are available in the versioned output replication_holm_sensitivity.csv. These post-outcome calculations did not alter the registered replication rule.

| Table S3. Continuous contrasts with BY-adjusted between-wave heterogeneity | | | | | | |
| --- | --- | --- | --- | --- | --- | --- |
| **ID** | **Exposure** | **2013 contrast** | **2019 contrast** | **2019-2013 difference (95% CI)** | **BY p** | **2013 discovery** |
| 4 | Self-rated health | 6.21 | 7.30 | 1.09 (0.57 to 1.60) | 0.002 | Yes |
| 7 | Self-rated oral health | 2.61 | 3.76 | 1.15 (0.67 to 1.64) | <0.001 | Yes |
| 27 | Weekly fruit-consumption frequency | -0.35 | -0.79 | -0.45 (-0.69 to -0.20) | 0.013 | Yes |
| 32 | Participation in religious activities | 0.21 | -0.15 | -0.36 (-0.59 to -0.14) | 0.048 | No |
| **Legend**. Contrasts are adjusted mean PHQ-9 score differences. No binary contrast met the BY-adjusted heterogeneity criterion. A nonsignificant test does not establish equivalence. All 31 between-wave contrasts are displayed in Figure S3. | | | | | | |

##### Missingness, internal consistency, and survey-design diagnostics

| Table S4. Exposure-wave rows with weighted complete-case availability below 99% | | | | | |
| --- | --- | --- | --- | --- | --- |
| **ID** | **Exposure** | **Wave** | **Exposure missing, n** | **Weighted complete** | **PHQ-9 >=10: complete - incomplete** |
| 11 | Hypertension | 2013 | 1787 | 97.0% | +5.0 pp |
| 33 | Household sanitation classification | 2013 | 2576 | 97.2% | +0.8 pp |
| 11 | Hypertension | 2019 | 1669 | 98.0% | +5.4 pp |
| **Legend**. pp = percentage points. Structural non-applicability is excluded from the denominator. No exposure-wave row had weighted complete-case availability below 95%. | | | | | |

| Table S5. Descriptive PHQ-9 internal consistency by wave | | | | |
| --- | --- | --- | --- | --- |
| **Wave** | **Complete PHQ-9, n** | **Weighted alpha** | **Weighted Pearson omega** | **Weighted item-rest range** |
| 2013 | 60202 | 0.845 | 0.855 | 0.405 to 0.679 |
| 2019 | 88531 | 0.859 | 0.868 | 0.423 to 0.701 |
| **Legend**. Omega is a one-factor approximation based on the survey-weighted Pearson correlation matrix; no design-based confidence interval was calculated. These estimates describe internal consistency, not test-retest reliability. | | | | |

Lonely-PSU diagnostic. ID 40 in PNS 2013 had 20 analytic strata with one contributing PSU. Repeating the binary and continuous models with survey.lonely.psu = 'average' reproduced the registered 'adjust' estimates and standard errors to numerical precision.

##### Exposure-shape diagnostics

| Table S6. Post-outcome ordinal nonlinearity diagnostic summary | | | | | |
| --- | --- | --- | --- | --- | --- |
| **Wave** | **Outcome** | **Applicable** | **Raw p<0.05** | **Holm p<0.05** | **Holm-significant exposure IDs** |
| 2013 | Binary PHQ-9 >=10 | 7 | 6 | 5 | 4, 7, 28, 46, 47 |
| 2019 | Binary PHQ-9 >=10 | 7 | 4 | 4 | 4, 28, 42, 47 |
| 2013 | Continuous PHQ-9 | 7 | 7 | 7 | 4, 7, 28, 33, 42, 46, 47 |
| 2019 | Continuous PHQ-9 | 7 | 7 | 7 | 4, 7, 28, 33, 42, 46, 47 |
| **Legend**. Holm correction was applied separately within each wave-by-outcome diagnostic family. ID 3 was retained in the protocol but was not applicable because it had two levels. These post-outcome diagnostics cannot alter registered classifications; nonsignificance is not proof of linearity. | | | | | |

| Table S7. Post-outcome discrete-score shape diagnostic summary | | | | | |
| --- | --- | --- | --- | --- | --- |
| **Wave** | **Outcome** | **Family** | **Tests** | **Holm p<0.05** | **Holm-significant IDs** |
| 2013 | Binary PHQ-9 >=10 | General | 8 | 2 | 27, 45 |
| 2019 | Binary PHQ-9 >=10 | General | 8 | 2 | 27, 32 |
| 2013 | Continuous PHQ-9 | General | 8 | 5 | 27, 30, 31, 32, 45 |
| 2019 | Continuous PHQ-9 | General | 8 | 5 | 27, 30, 31, 32, 45 |
| 2013 | Binary PHQ-9 >=10 | Occupational | 1 | 1 | 38 |
| 2019 | Binary PHQ-9 >=10 | Occupational | 1 | 1 | 38 |
| **Legend**. The 34 tests are post-outcome diagnostics. Holm correction was applied separately within each wave-by-outcome-by-family set. Nonsignificance is not proof of linearity, and significance cannot alter registered effect estimates, discoveries, or replications. Flexible direct contrasts for the three primary binary discoveries modeled as scores retained their direction and similar magnitude; full estimates are in the OSF output registered_vs_flexible_target_contrasts.csv. | | | | | |

| Table S8. Support for the occupational harmful-agent count (ID 38) | | | | | |
| --- | --- | --- | --- | --- | --- |
| **Wave** | **Score** | **n** | **PHQ-9 positive n** | **Weighted share, %** | **Weighted PHQ-9 positive, % (SE)** |
| 2013 | 0 | 15,822 | 882 | 42.568 | 4.75 (0.28) |
| 2013 | 1 | 10,663 | 761 | 28.510 | 6.16 (0.35) |
| 2013 | 2 | 5,902 | 469 | 17.375 | 7.61 (0.62) |
| 2013 | 3 | 2,765 | 252 | 8.010 | 8.24 (0.82) |
| 2013 | 4 | 974 | 96 | 2.687 | 8.45 (1.26) |
| 2013 | 5 | 245 | 23 | 0.692 | 8.46 (2.78) |
| 2013 | 6 | 47 | 2 | 0.119 | 0.47 (0.43) |
| 2013 | 7 | 14 | 4 | 0.039 | 64.20 (20.50) |
| 2019 | 0 | 26,944 | 2,196 | 50.962 | 8.39 (0.31) |
| 2019 | 1 | 13,386 | 1,186 | 24.510 | 9.70 (0.51) |
| 2019 | 2 | 7,033 | 604 | 14.166 | 8.87 (0.65) |
| 2019 | 3 | 3,521 | 301 | 7.194 | 7.02 (0.69) |
| 2019 | 4 | 1,153 | 145 | 2.325 | 12.35 (1.57) |
| 2019 | 5 | 324 | 43 | 0.643 | 11.93 (2.81) |
| 2019 | 6 | 64 | 6 | 0.122 | 8.94 (4.59) |
| 2019 | 7 | 22 | 4 | 0.078 | 5.39 (3.78) |
| **Legend.** Counts are unweighted; percentages and standard errors are design-weighted, unadjusted support diagnostics. Scores 6-7 together represented 0.158% of the weighted 2013 domain and 0.200% of the weighted 2019 domain. Score 7 contained four PHQ-9-positive observations in each wave. These sparse endpoints do not support interpreting the registered slope-rescaled seven-versus-zero PR as a direct empirical contrast or linear dose response.  The separate occupational descriptive-family estimates are displayed in Figure S4; no occupational replication proportion was prespecified. | | | | | |

### Supplementary Figures

#### Figure S1. Secondary continuous PHQ-9 results across 31 general exposures.

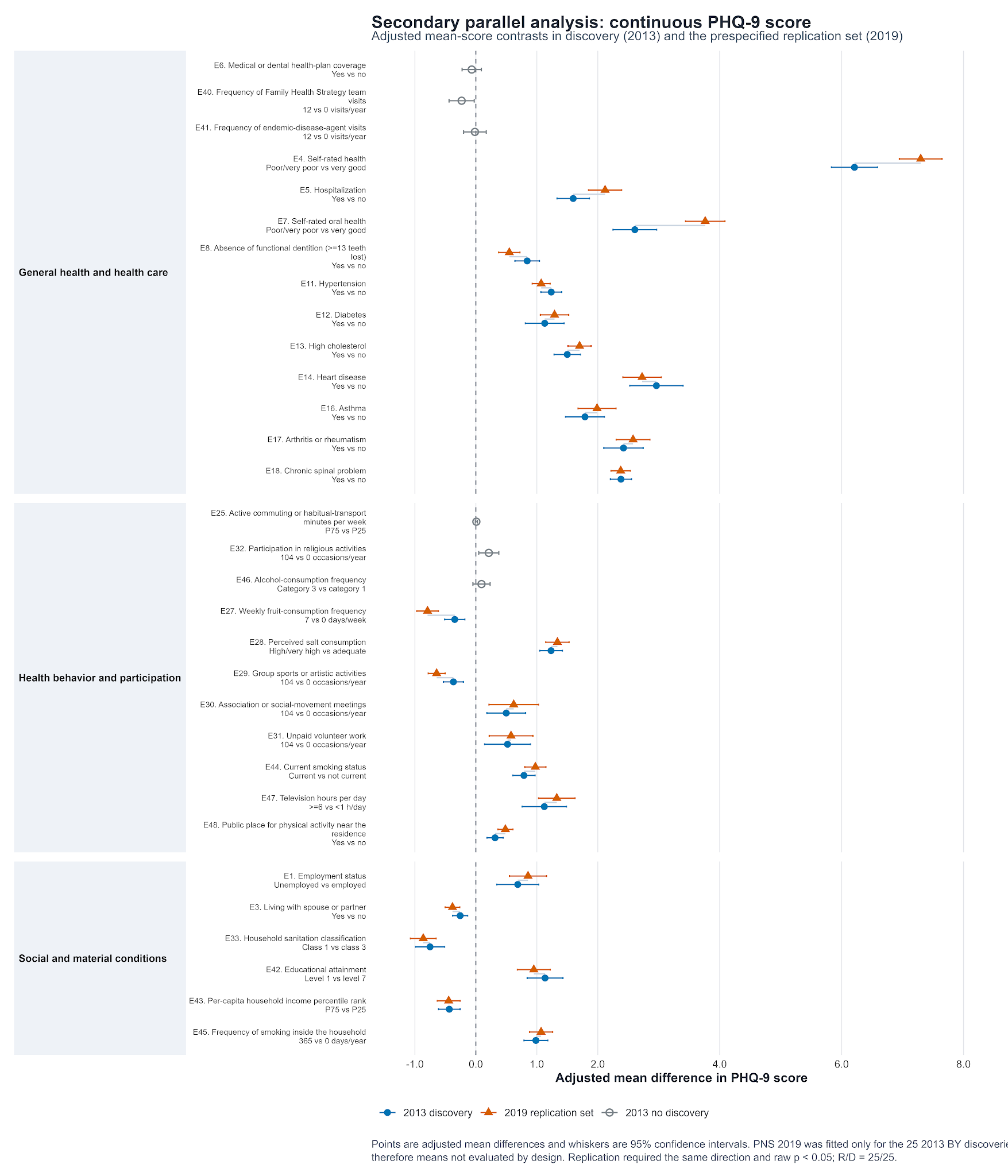

**Legend**. Points show adjusted mean-score differences and 95% confidence intervals. PNS 2019 estimates were fitted only for the 25 PNS 2013 BY discoveries.

#### Figure S2. Preregistered discovery-replication architecture. Solid boxes denote the frozen pre-outcome analysis; the dashed box contains post-outcome sensitivity analyses.

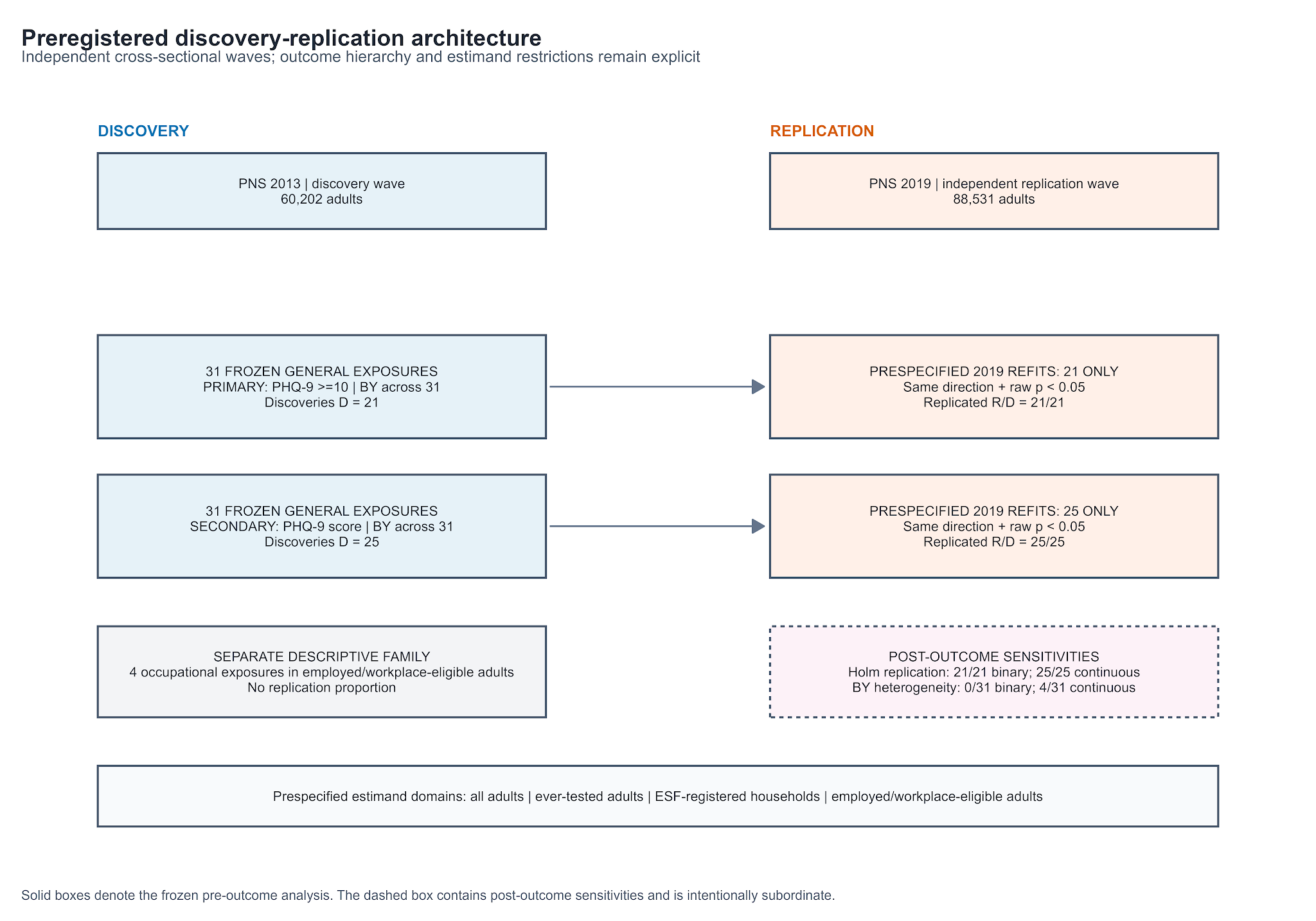

**Legend**. Solid boxes denote the frozen pre-outcome analysis. The dashed box contains post-outcome sensitivities and is intentionally subordinate.

#### Figure S3. Post-outcome between-wave heterogeneity sensitivity across all 31 general exposures.

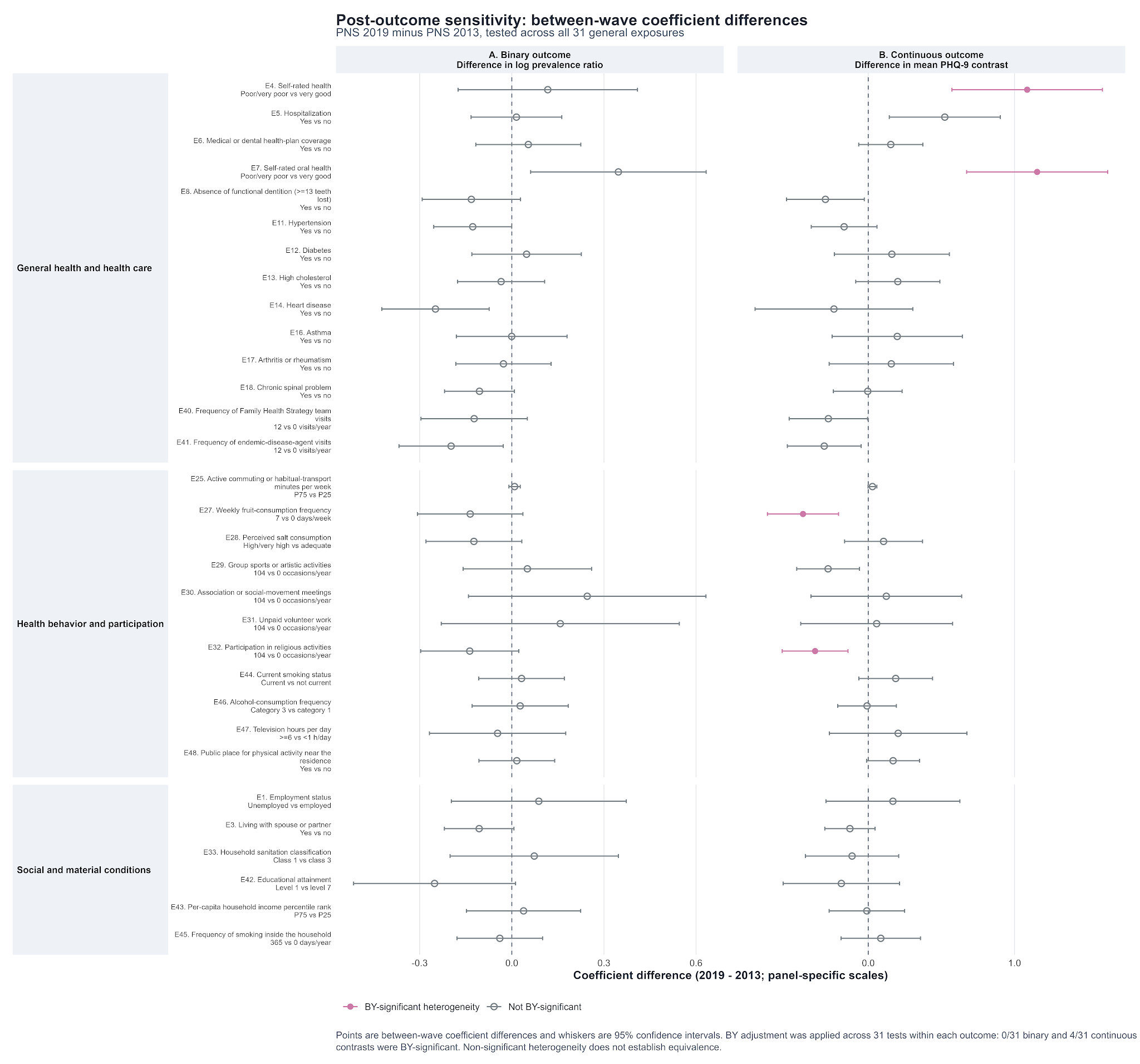

**Legend**. Differences are PNS 2019 minus PNS 2013; BY adjustment was applied separately by outcome. Nonsignificant heterogeneity does not establish equivalence.

#### Figure S4. Occupational descriptive-family results in employed or workplace-eligible adults.

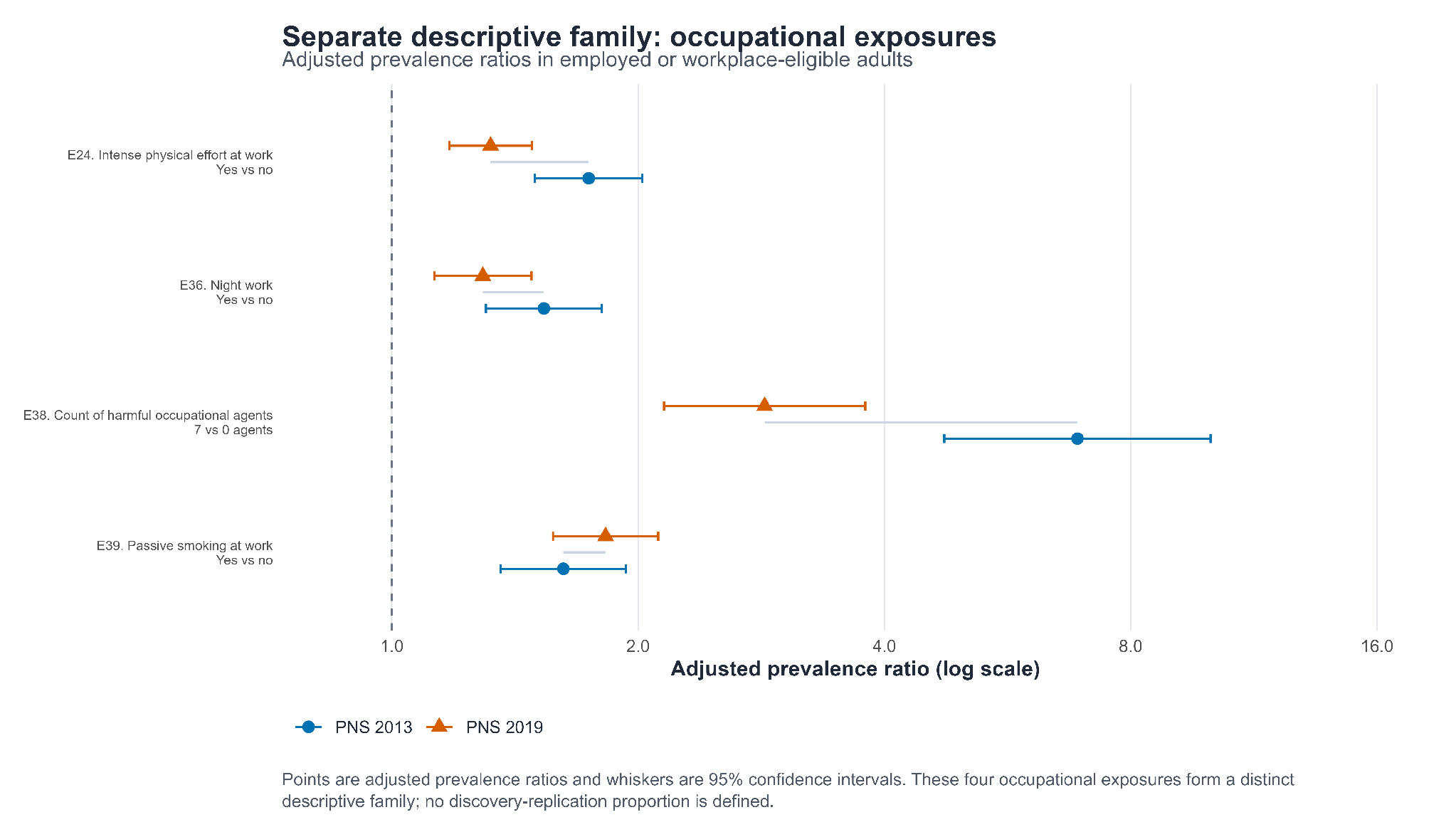

**Legend**. Points show adjusted prevalence ratios and 95% confidence intervals. The ID 38 seven-versus-zero harmful-agent-count effect is a model-implied rescaled slope; the other three effects are direct binary contrasts. No occupational replication proportion was prespecified.
